# Prevalence of mask wearing in northern Vermont in response to SARS-CoV-2

**DOI:** 10.1101/2020.07.23.20158980

**Authors:** Brian Beckage, Thomas E. Buckley, Maegan E. Beckage

**Affiliations:** Department of Plant Biology, University of Vermont, Burlington, Vermont, USA; Department of Computer Science, University of Vermont, Burlington, Vermont, USA; Gund Institute for Environment, University of Vermont, Burlington, Vermont, USA; Colchester High School, Colchester, Vermont, USA; Essex High School, Essex Junction, Vermont, USA

**Author notes:** These authors contributed equally to this work (Thomas E. Buckley, Maegan E. Beckage).

## Abstract

**Objectives:** Information on prevalence of face mask usage in response to SARS-CoV-2 is required to both model disease spread and to improve compliance with mask usage. We sought to (1) estimate the prevalence of mask usage in northern Vermont and to (2) assess the effect of age and sex on mask usage.

**Methods:** We monitored the entrances to businesses and visually assessed individuals’ age, sex, and mask usage from a distance. We collected 1004 observations from 16 May through 30 May 2020 as businesses began to reopen following an extended state-wide lockdown. We analyzed these data using Bayesian random effects logistic regression.

**Results:** 75.5% of individuals used a mask with significant effects of age and sex on mask usage. Females were more likely than males to wear masks (83.8%, n=488 vs. 67.6%, n=516); the odds of mask usage in males were 53% of those for females. Elders were most likely to wear a mask (91.4%, n=209) followed by young adults (74.8%, n=246), middle-aged adults (70.7%, n=519) and children (53.3%, n=30). The odds of an elder wearing a mask were 16.7 times that of a child, while the odds for young adults and middle-aged adults were ∼3 times greater than for a child. Highest mask usage was in elder females (96.3%, n=109) and lowest mask usage was in male children (43.8%, n=16).

**Conclusions:** We found high prevalence of mask usage overall, but also large differences in mask usage with age and sex. Females and elders had the highest use of face masks.

## Introduction

The use of face masks can be effective at reducing the transmission of SARS-CoV-2^1, 2^. Theoretical studies show that the extent of disease outbreaks is dependent on population compliance with mask usage and effectiveness of the masks at reducing viral transmission^3^. Simulation studies also suggest that there is a rapid phase transition from low to high prevalence of infection in populations as a function of compliance with social mitigation strategies such as mask usage. Mask usage below a threshold percentage of individuals results in widespread infection of the population^4^. Information on population compliance with mask usage is thus important to understanding the continuing dynamics of SARS-CoV-2.

Our objective in this study was i) to estimate the prevalence of mask usage in public places of business and ii) to assess the effect of sex and age on mask usage in northern Vermont. We visually assessed mask usage and estimated age and sex of individuals from a distance without any interaction with the patrons. We collected 1004 observations from outside 8 businesses and analyzed these data using a logistic regression models.

## Materials and methods

### Regional context

Vermont reported its first case of SARS-CoV-2 on 7 March 2020. The governor declared a state of emergency on 13 Mar 2020, followed by a series of orders to mitigate spread of SARS-CoV-2^5^. These included a “Stay Home, Stay Safe” order on 24 March that directed the closure of in-person operations for all non-essential businesses and mandated that the public remain at home unless leaving for reasons critical to health or safety. On 15 May 2020, the order was updated to “Be Smart, Stay Safe”, relaxing the previous restrictions and beginning the process of reopening businesses, while urging Vermonters to socially distance and use masks in public.

Our study was carried out in Chittenden County, located in northwestern Vermont, USA. Chittenden County is the most urbanized and densely populated county in Vermont, containing 26.2% of the state population; population estimates and demographics for Chittenden County are available from the US Census Bureau^6^.

### Data collection

We assessed the prevalence of mask wearing by passively observing the entrances to businesses from 16 May through 30 May 2020 that included grocery, hardware, and convenience stores, and a golf course. We recorded individuals’ mask usage as well as their estimated age and sex. Age and sex were assessed visually without interactions with the individuals. We assigned age into the following categories: child (≤14 years old), young adult (>14 and ≤25 years old), middle age adult (>25 and ≤60 years old) and elderly adult (>60 years old). Mask use was determined as individuals were entering the establishment since they sometimes carried the mask in their hand in the parking lot and only put on masks when entering the store. The observers were unobtrusively stationed in a vehicle near the store entrance or outside the businesses and did not interact with the observed individuals. The University of Vermont Research Protections Office granted our study an exemption from review by our institutional review board (CHRBSS (Behavioral): STUDY00001094).

### Statistical analysis

We analyzed these data using logistic regression. We first selected the best-supported model of mask usage from a set of fixed-effect logistic regression models using Akaike’s Information Criterion^7^. These models were fit using the GLM function in R^8^. We then added business type as a random effect to the selected model and fit this Bayesian random-effects logistic regression using the NIMBLE library within R^9^. We did not treat business locations as the random effect but rather the business identity, e.g., all supermarkets of a given chain were treated as a single random effect. This implies that any effect of a given business is associated with the demographic utilizing that business brand (e.g., grocery store chain 1 vs. grocery store chain 2) as opposed to an effect of any particular store location. We ran 4 MCMC chains, each of length 2 million, used a burn-in length of 10,000, and thinned our chains by a factor of 100. We used Gelman and Rubin’s Convergence Diagnostic to confirm convergence of our chains^10^. We report parameter estimates and odds ratios.

## Results

Mask usage was 75.5% overall, but varied widely with sex and age group (Table 1). Mask usage for males was 67.6% compared to 83.8% for females. Mask use was highest for the elders age group with 91.4% mask use. Children had the lowest rate of compliance at 53.3%, though our sample size of children was low compared to other groups. Highest overall use of face masks was elder females with 96.3% compliance, while lowest compliance was with male children (43.8% mask use), followed by middle age males (62.5% mask use).

**Table 1.** Number of people wearing mask/not wearing mask (and % mask use) by age and sex for a total of 1004 observations.

|  | Male | Female | Total |
| --- | --- | --- | --- |
| Children | 7/9 (43.8 ) | 9/5 (64.3) | 16/14 (53.3) |
| Young adults | 78/37 (67.8) | 106/25 (80.9) | 184/62 (74.8) |
| Middle-aged | 178/107 (62.5) | 189/45 (80.8) | 367/152 (70.7) |
| Elders | 86/14 (86.0) | 105/4 (96.3) | 191/18 (91.4) |
| Total | 349/167 (67.6) | 409/79 (83.8) | 758/246 (75.5) |

The most parsimonious, fixed effects model of mask usage included age and sex but not their interaction (Table 2). The sex:age interaction term was, however, included in the second ‘best’ fixed effects model. Our final model, which included age and sex as fixed effects, and business type as a random effect, estimated that the odds of a male using a mask were only 53% of those of a female (Table 3). The odds of an elder wearing a mask were >16 times greater than a child and ∼5 times (5.59 and 5.12, respectively) that of young or middle-aged adults, while the odds for young adults and middle-aged adults were ∼3 times (2.99 and 3.26, respectively) greater than for a child.

**Table 2.** Comparison of fixed effects models using AIC.

| Model | AIC | $\Delta$ AIC |
| --- | --- | --- |
| 1. Intercept only | 1120.05 | 74.63 |
| 2. Sex | 1085.89 | 40.47 |
| 3. Age | 1077.57 | 32.15 |
| 4. Sex + Age | 1045.42 | 0 |
| 5. Sex + Age + Sex:Age | 1049.98 | 4.56 |

**Table 3.** Parameter estimates of fixed sex and age effects for final model.

|  | Mean | 2.5% | 50% | 97.5% | Odds |
| --- | --- | --- | --- | --- | --- |
| Intercept | -0.24 | -2.73 | -0.22 | 2.11 | – |
| Male | -0.65 | -0.99 | -0.65 | -0.31 | 0.53 |
| Young | 1.00 | 0.15 | 1.00 | 1.85 | 2.99 |
| Middle Age | 1.10 | 0.28 | 1.10 | 1.91 | 3.26 |
| Elder | 2.69 | 1.72 | 2.69 | 3.68 | 16.69 |

There was large variation in mask usage across businesses identities (Fig 1). The lowest mask usage was at a golf course (LF), likely because the individuals were outdoors and thus perceived less need to wear a mask, followed by a gas station convenience store (MF) (Table 4). The highest mask usage was associated with a grocery store (SH).

**Table 4.**
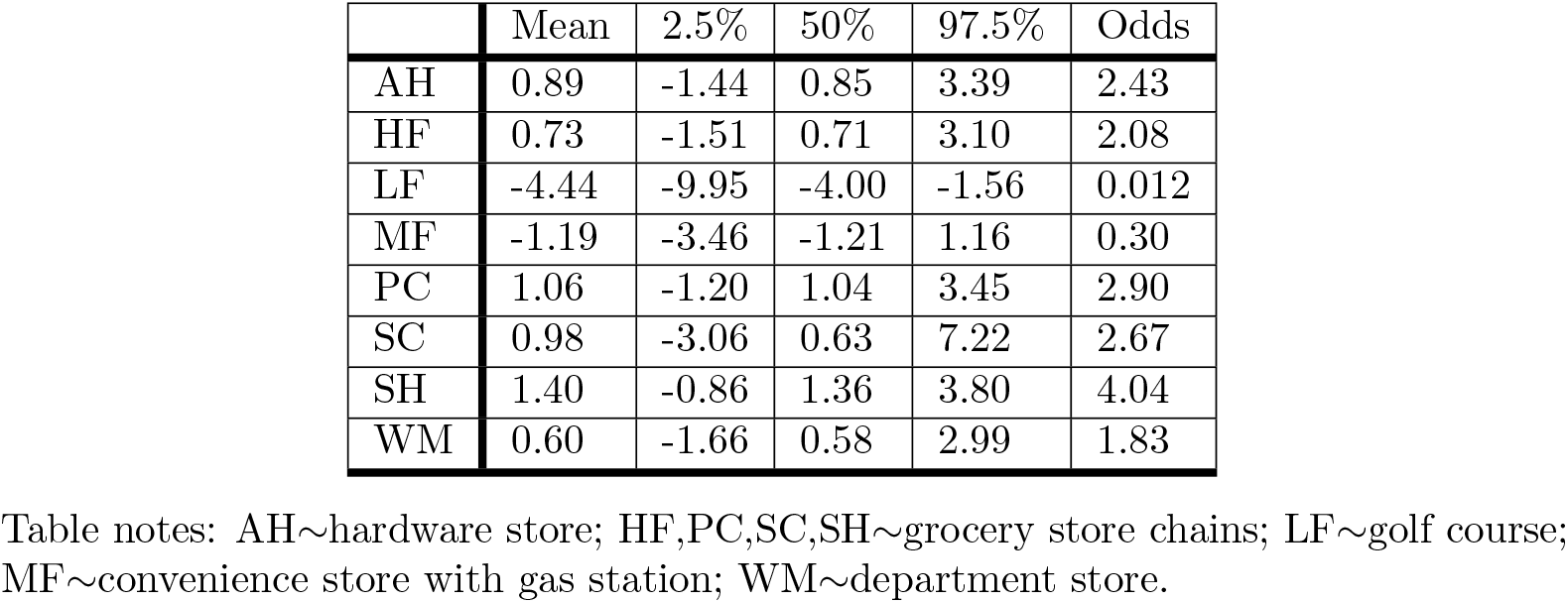
Estimates of random effects of eight business types in final model.

**Fig 1.**
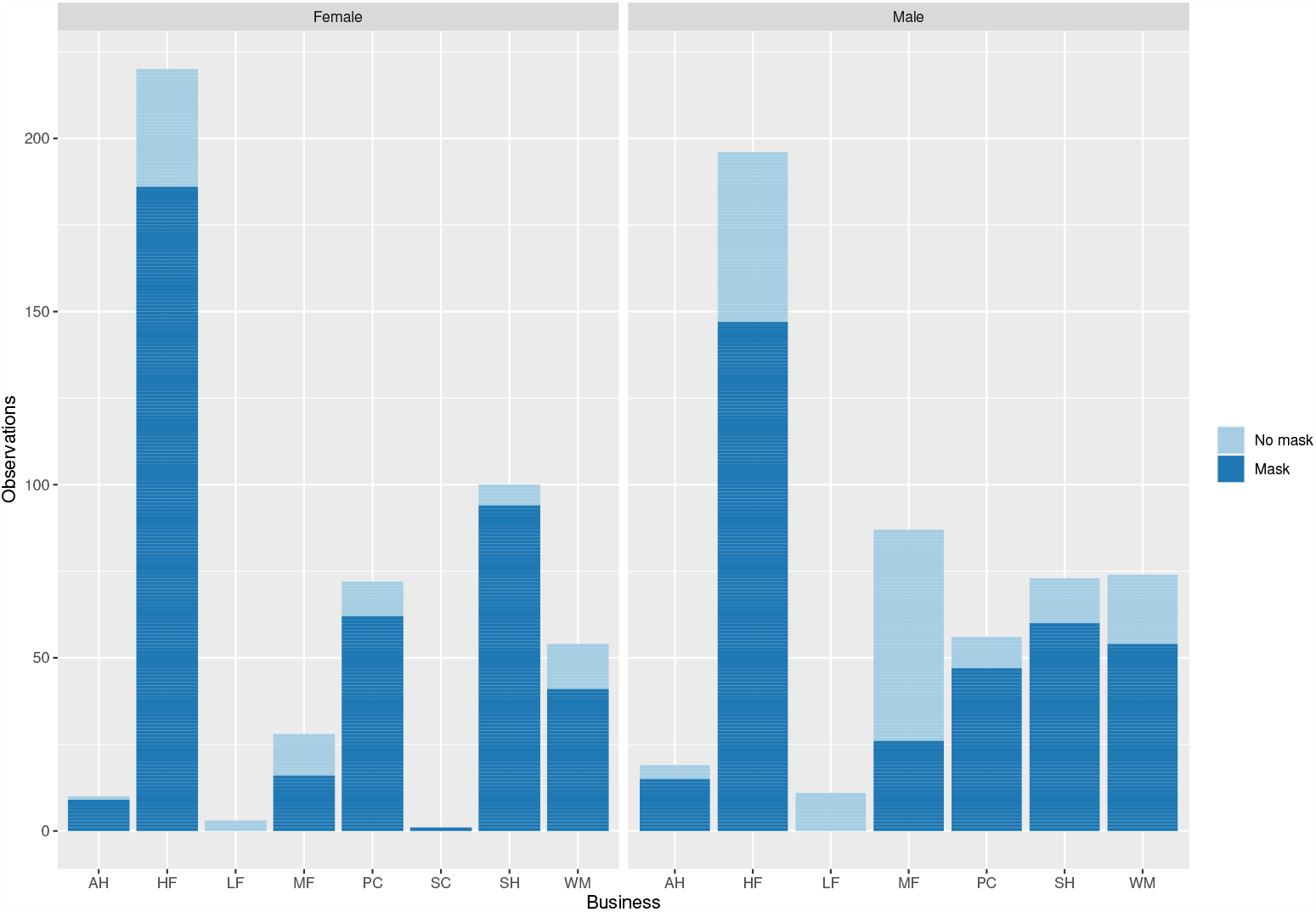
Mask usage as a function of sex and business type. Business types are defined in Table 4.

## Discussion

The overall mask usage of 75.5% found in our study was high compared to the 41.2% mask usage observed in grocery stores in Wisconsin over the same period^11^. Similar trends to what we observed with respect to sex and age were also found in their study: Mask usage was higher in older adults (59.5%) and females compared to males (44.8% vs 36.9%, respectively), and lowest mask usage was among minors (26.2%). We do note that infections have continued to increase in Wisconsin^12^ compared to Vermont^13^, although this is just an observation rather than a claim that these divergent trends are related to differences in mask usage.

## Conclusion

Social mitigation strategies such as wearing masks in public are important to reducing spread of SARS-CoV-2. Information on compliance with use of face masks is required both for constructing models of disease spread and also for targeted messaging to increase compliance with mask usage. Studies such as this provide baseline information for assessing both demographic effects on mask usage, and also provide baseline information for regional comparisons or to assess trends in mask usage across time within a region.

## Data Availability

We have posted the data and code to a GitHub repository and will make these available publicly after the study has been published in a peer-reviewed journal.

https://github.com/brianbeckage/Masks_covid19.git

## Acknowledgments

We acknowledge Prof. Jane Molofsky for reading and commenting on an earlier draft of this manuscript.

